# Two Billion Infected: An Inexpensive Method to Measure Latent Toxoplasmosis and its Economic Consequences^*^

**DOI:** 10.1101/2024.07.27.24311112

**Authors:** Anja Achtziger, Carlos Alós-Ferrer, Michele Garagnani

## Abstract

Over two billion people worldwide are infected with the protozoan Toxoplasma gondii, which influences human behavior and cognition. Standard diagnosis methods involve costly medical tests, which prevents widespread testing and hinders the study of the infection’s effects. We propose and validate an inexpensive and easy-to-deploy diagnostic method for latent Toxoplasmosis infections using response times and finite-mixture models. A clinical study showed that the method is sensitive and accurate. A large, representative UK study showed that the infection’s consequences are relevant and pervasive, both economically (decrease in yearly income and employment) and behaviorally (increase in risky behaviors, stress, and depression).

---

The protozoan Toxoplasma gondii causes a worldwide parasitic infection for which both the CDC and the WHO consider it a priority to improve diagnostic testing (CDC, 2020). Evidence of exposure to this parasite, which is transmitted from felines to humans, is found in approximately 30% of the world’s human population, i.e. around 2.4 billion people (Montoya and Liesenfeld, 2004). The latent Toxoplasmosis infection has been linked to behavioural alterations in humans, ranging from increased suicide rates and traffic accidents (Kocazeybek et al., 2009; Milne et al., 2020) to increased risk of psychiatric disorders and increased impulsivity (Sutterland et al., 2015, 2020). Current diagnosis methods for this infection rely on relatively expensive and time-consuming medical tests, e.g., saliva, blood samples, or autopsical analyses (Saadatnia and Golkar, 2012). This has prevented widespread testing, especially in less-than-wealthy regions, and in particular has hindered the systematic study of the socioeconomic and behavioral effects of latent Toxoplasmosis infections.

We propose and validate an inexpensive and easy-to-deploy method to test for the prevalence of Toxoplasmosis in the population, and use it to study the behavioral and economic effects of the latent infection. The results of a clinical trial and a large, representative-sample study strongly support the validity of the proposed method. While the method cannot and does not aim to substitute medical tests, it can be used for large-scale testing to understand the infection’s behavioral and economic consequences, as well as to inform and design policies aimed at reducing, preventing, and eventually eradicating infections in the population. Our results uncover previously-undocumented behavioral, economic, and psychological effects of latent Toxoplasmosis and confirm previously-hypothesized effects. We find relevant and pervasive effects on economic outcomes and economic behavior, including a decrease in yearly income (2500 GBP), an increase in unemployment (of around 11%), a decrease in socioeconomic status, increased risk-taking and reduced patience (according to the survey measures of Falk et al., 2018), reduced self-control, and increased participation in risky economic initiatives (startups, entrepreneurship). We also find clear evidence of negative effects on other risky behaviors (e.g., increased alcohol consumption and smoking) and mental health issues (depression, anxiety, stress, and mental disorders in general).

Our objective is to enable inexpensive, large-scale studies on the behavioral and economic consequences of the infection. The method we propose, however, can also be seen as a diagnostic tool for latent Toxoplasmosis infections. While less clinically relevant than the acute condition, latent infections are still of primary importance as they are characterized by low-grade persistent neuroinflammation and cardiovascular injury (Egorov et al., 2021), can act as a reservoir for acute-stage reactivation causing disease in immunocompromised patients and might exacerbate the extent of brain damage after traumatic brain injury (Robert-Gangneux and Dardé, 2012; Baker et al., 2024). Crucially, about three-quarters of new infections in healthy individuals are asymptomatic (Weiss and Dubey, 2009). Hence, a cheap and easy-to-deploy method to diagnose latent Toxoplasmosis infections would increase our ability to prevent further complications for the affected population.

A number of behavioral changes in Toxoplasma-infected individuals are well-established (Milne et al., 2020). Further, differences between Toxoplasma-infected and Toxoplasma-free subjects usually increase with time since infection and the intensity of changes correlates with the level of Toxoplasma IgG antibodies (Flegr and Havlíček, 2013). Behavioral changes are usually interpreted as the product of the parasite’s manipulative ability (Mehlhorn, 2015; Houdek, 2017). The parasite can synthesize tyrosine hydrolase (an enzyme involved in dopamine biosynthesis) which contributes to dopamine disregulation among the infected (McConkey et al., 2013). Toxoplasma gondii also alters the expression of a range of other neurotransmitters, including *γ*-Aminobutyric acid (GABA), glutamate, serotonin, and norepinephrine (Chaudhury and Ramana, 2019). Changes in dopamine-mediated neurotransmission have been proposed to be involved in addictive and obsessive behaviours (Kalivas and Volkow, 2005), and serotonin is thought to play a central role in the etiology of mood disorders (Kishi et al., 2013).

The method we propose is based on the physiological changes induced by the parasite. In particular, latent Toxoplasmosis is known to cause longer response times in part of the population, and response times are easy to measure even in the field. Specifically, people with RhD-negative blood type become slower when afflicted by Toxoplasmosis, while RhD-positive individuals do not experience a change in the distribution of response times (Havlíček et al., 2001; Novotná et al., 2008). We leverage the statistical properties of the distribution of response times to reliably estimate the distribution of the latent infection in the RhD-negative subjects using finite-mixture models, resulting in a classification in imputed-afflicted and imputed-nonafflicted which can be used to test for behavioral and economic effects in large samples without relying on expensive medical tests. As long as one can assume that the probability of Toxoplasmosis infection is independent of the blood type, the proposed method also delivers an estimate of the prevalence of the infection in the general population.

Figure 1 describes our method and illustrates it with data from our clinical-trial study (see Figure 3 for the corresponding illustration for our survey study). Response times (RTs) are known to be log-normally distributed (Spiliopoulos and Ortmann, 2018), i.e. log-transformed response times are normally distributed. Suppose that a given population is split into two groups, the afflicted and the non-afflicted. If the afflicted show a systematic delay in RTs, then their log-transformed RT distribution is shifted compared to the non-afflicted. This is illustrated in Figure 1(top-left). The distribution for the entire population is then a *mixture*, i.e. part of the population follows one distribution and the rest follows a different distribution. While a linear combination of normal distributions is always normal, mixtures of normals with different means are generally not normal. On the contrary, they can become bimodal depending on the relative size of the groups (Figure 1, top-right). In our clinical-trial study, data shows this characteristic pattern for RhD-negatives but not for RhD-positives (Figure 1, bottom-left), in line with the predicted physiological changes due to the infection. Mixtures of normally-distributed variables, however, are identifiable through appropriate statistical methods, in particular finite-mixture models. Applying these models, we can recover the characteristics of the two groups, i.e., the two distributions and their relative sizes (Figure 1, bottom-right).

**Figure 1:**
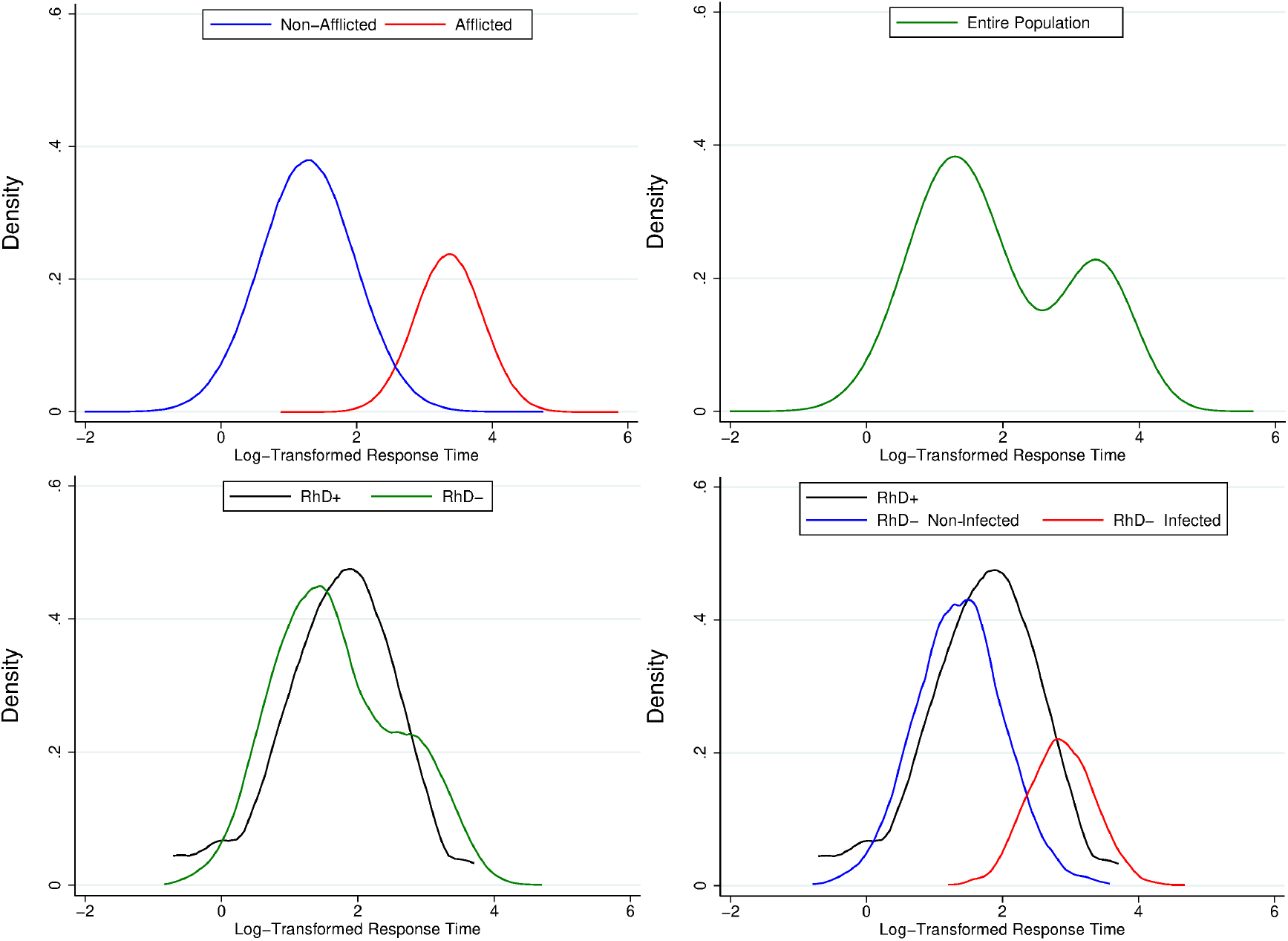
If afflicted individuals in a population show a systematic reaction slow-down, the distribution of their log-transformed response times is shifted compared to the nonafflicted (top-left panel). The distribution for the entire population is then a mixture showing a bimodal pattern (top-right panel). Consistent with the expected effects of Toxoplasmosis infection, we find this pattern in our clinical study for RhD-negative individuals, but not for RhD-positives (bottom-left panel). Applying finite-mixture models to the data, we recover the subgroup distributions and their relative sizes (bottom-right panel). This delivers an estimate of the extent of the infection in the population and also individual-level infection probabilities.

Crucially, the finite-mixture model estimates the probability that each individual belongs to one or the other subgroup. Hence, our approach can classify an individual as potentially having a latent Toxoplasmosis infection or not using only response times, delivering a non-clinical test at the individual level. To validate the proposed method (for the purposes of studying behavioral and socioeconomic effects in large samples), in our clinical trial we used this classification in comparison with a standard blood test, which verifies the presence of the corresponding IgG antibodies. We show that our new method displays a good performance compared to the more-expensive medical procedure, while being much easier to deploy on a larger scale.

The purpose of our first, clinical trial, was to establish the validity and accuracy of the method. For this reason, that trial included an antibodies test for Toxoplasmosis. In contrast, the purpose of our second study was to illustrate the usefulness and ease of application of the method in large (online) samples, where direct tests requiring blood collection are expensive and impractical. Hence, our second study used our method, as validated in the first study, to examine the economic and behavioral consequences of Toxoplasmosis by conducting a survey measuring RTs with a large, representative sample of the UK population.

Even though the survey could not include direct medical tests, we also obtain additional validations by replicating and confirming behavioral results previously reported in the literature for infected individuals, using our RT-based classification instead. More importantly, we also identify new insights on the pervasive effects of the latent infection. We show that the infection has sizable socioeconomic consequences. The group imputed to be infected reports higher unemployment (by around 11%), lower perceived social ranking, and a lower annual income (of about 2,500 GBP on average). Further insights also include increased risk-seeking and impatience as captured by the survey measures of Falk et al. (2018), problematic behaviors as reduced self-control, increased sensation-seeking, and increased alcohol consumption, and also mental-health issues including depression, anxiety, and stress.

## I Experimental Procedures and Participants

We conducted two studies: a clinical study and a survey study. All hypotheses, sample size, and data analysis plans for both studies were preregistered at ClinicalTrials.gov, NCT05860998 (clinical study) and AsPredicted.org, study number 108329 (survey study; https://aspredicted.org/WS4_34R). The studies were conducted according with Swiss law and internationally-recognized principles (Declaration of Helsinki). They received institutional review and approval from the Human Subjects Committee of the Faculty of Economics of the University of Zurich (OEC-IRB-2022-077) and the Ethics Committee in the Canton of Zurich (BASEC:2023-00366). Raw data files for each study and the code for the analyses are available on OSF https://osf.io/j2rku/?view_only=482ca01c948044a08f5a8244172d4090. No observation was excluded from the analyses.

The clinical study was a controlled trial performed at the Laboratory for Social and Neural Systems Research (SNS) at the University Hospital of Zurich. We recruited *N* = 119 participants (51.28% females; mean age 23.83, SD 2.93). Participants were informed about the purpose of the study only at the end. During recruitment, they were only informed about its general design, inclusion criteria, and the fact that a blood sample would be required. Inclusion criteria were age between 18 and 35 years, willingness to participate in the study, written declaration of consent, good English language skills, and reporting RhD-negative blood type. Criteria for exclusion were an inability to give informed consent, any neurological disorders, reduced general health or chronic diseases (e.g., autoimmune disease, severe cardiovascular diseases, insulin-dependent diabetes, and chronic pain disorders). Participants were made aware of these criteria when they were recruited and reminded on the day of the study. During the study, they were seated in front of a computer screen and performed a series of tasks (described below). After completing these tasks, participants moved to another room with a hospital cot, where a nurse drew their blood. Participants were paid a flat fee (equivalent to 25 GBP at the time of the study), and the average duration for the study was 30 minutes.

Blood samples were tested for the presence of Toxoplasma gondii IgG antibodies. We further tested whether the reported blood type was correct with the blood sample (40 participants actually had RhD-positive blood type). The analyses of the blood samples were performed by Medica, Medizinische Laboratorien, Dr. F. Kaeppeli, Wolfbachstrasse 17, 8024 Zurich. We informed participants about the result of the tests when they were available.^1^

The second study involved a representative sample of the UK population and was performed on Prolific, an online recruitment system (Palan and Schitter, 2018). When registering to Prolific participants need to provide demographic and socioeconomic information, which is delivered to researchers in an anonymized form. Prolific had added a question on blood types (both group and Rh factor) at our request several weeks before the start of the study. This allowed us to recruit RhD-negative and RhD-positive participants in equal and sufficient numbers (the sample is representative conditional on the blood type). Moreover, this made the participants completely blind to the aim of the study. Inclusion criteria for the survey were having reported their blood type to Prolific and completing the study, and passing a standard attention check at the beginning of the survey. We recruited *N* = 2, 020 participants (66.49% females, mean age 42.06, SD 13.06).^2^ Participants were paid a flat fee of 5 GBP for an average participation of 20 minutes. They were unaware of the research question and performed the same tasks as in the first study.

## II Method Validation: Clinical Trial

The main purpose of the clinical-trial study was to test whether the proposed method is able to predict infections assuming the medical test to be the ground truth. This creates a contingency table which can be tested using a *χ*^2^ test. The sample size to guarantee an a priori sufficient power (0.8) for a standard significance level (*α* = 0.05) with a medium effect size (*w* = 0.15) is *N* = 69. Because we could not tell in advance how many participants would actually be RhD-negative, we conservatively set to collect *N* = 120 independent observations.

Between July 3rd and November 8th 2023, *N* = 119 subjects (there was one no-show) participated in a clinical trial testing whether the proposed method is able to accurately detect latent Toxoplasmosis infections using only response times. For this purpose, blood samples were collected from all participants and a Toxoplasma gondii IgG Antibodies test was conducted. Of the 119, 79 (66.39%) actually were RhD-negative, as required for the analysis since the Toxoplasma-induced delay in RTs is only observable for this blood type (Havlíček et al., 2001; Novotná et al., 2008).

Among the 79 RhD-negative participants, 12 (15.19%) tested positive for a latent Toxoplasmosis infection according to the blood test. Our method classifies 13 (16.46%) of the RhD-negative participants as likely to have a latent Toxoplasmosis infection (i.e., more likely to belong to the group with longer RTs). The accuracy of the new method can then be directly tested using a *χ*^2^ test, which is highly significant (Pearson’s *χ*^2^ = 58.219, *p* < 0.0001, large effect size *w* = 0.85). As Figure 2(left) shows, the proposed method is sensitive (91.66%), as it classifies 11 of the 12 Toxoplasmosis-infected participants correctly, and specific (97.01%), as it only falsely classifies 2 of the 67 Toxoplasmosis-free participants.

**Figure 2:**
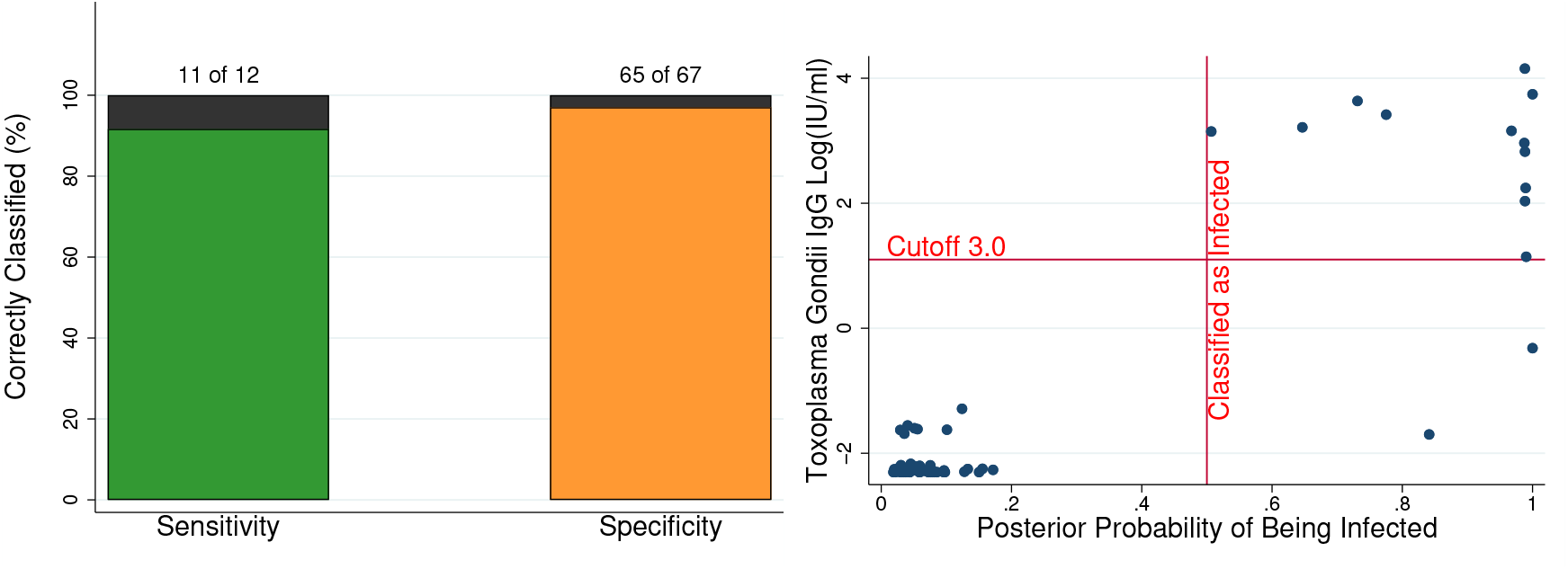
Left-hand panel: The proposed method, which uses only response times and is cheaper and easier to deploy at large scale than antibodies tests, presents high sensitivity (91.66%) and specificity (97.01%). Right-hand panel: The posterior probability of each participant being classified as infected by the proposed method is highly correlated with the individual proportion of Toxoplasma gondii IgG antibodies (log-transformed for ease of representation, Pearson’s *ρ* = 0.7125, *p* < 0.0001).

Since the blood test is based on a threshold value for antibody concentration, we also investigate the relation between the continuous measures underlying the variables above. We compare the posterior probability of being classified as infected by the proposed method with the proportion of Toxoplasma gondii IgG antibodies in participant’s samples (Figure 2(right)). We find a strong correlation (Pearson’s *ρ* = 0.7125, *p* < 0.0001) between the two measures, further attesting that the proposed method agrees with the antibodies test.

## III New Insights: Survey Study

To study the socioeconomic and psychological effects of Toxoplasmosis in a large sample, we included the reaction-time task (Havlíček et al., 2001; Novotná et al., 2008) in a survey of the UK population (*N* = 2, 020) with 50% RhD-negative subjects, conducted between the 3^*rd*^ and the 13^*th*^ of October 2022. The aim was to test for differences in economic outcomes and behavior between the groups imputed to be Toxoplasma-infected and Toxoplasma-free within the RhD-negative group according to non-parametric tests (Mann-Whitney U test). Because afflicted and non-afflicted are not equally numerous this decreases the power of the test. We conservatively established the sample size to have enough power (0.8) to detect a small effect size (*d* = 0.2). The required sample size is then *N* = 1, 010 participants, for a total of *N* = 2, 020 counting both RhD-negative and RhD-positive individuals. The RhD-positive group is then used as benchmark to ensure that the classification based on the proposed method or the blood types does not create artificial statistical differences.

The sample was otherwise representative of the overall population based on age and income. We followed the same procedure as in the clinical trial to recover the distributions of log-transformed RTs for imputed-infected and imputed-non-infected RhD-negative participants (Figure 3). The method classifies 185 participants (18.32% of the 1, 010 RhD-negatives) as infected. For comparison, the National Institute of Health estimates the incidence of Toxoplasmosis to be between 23% and 33% in the UK, depending on regional differences (Joynson, 1992).

**Figure 3:**
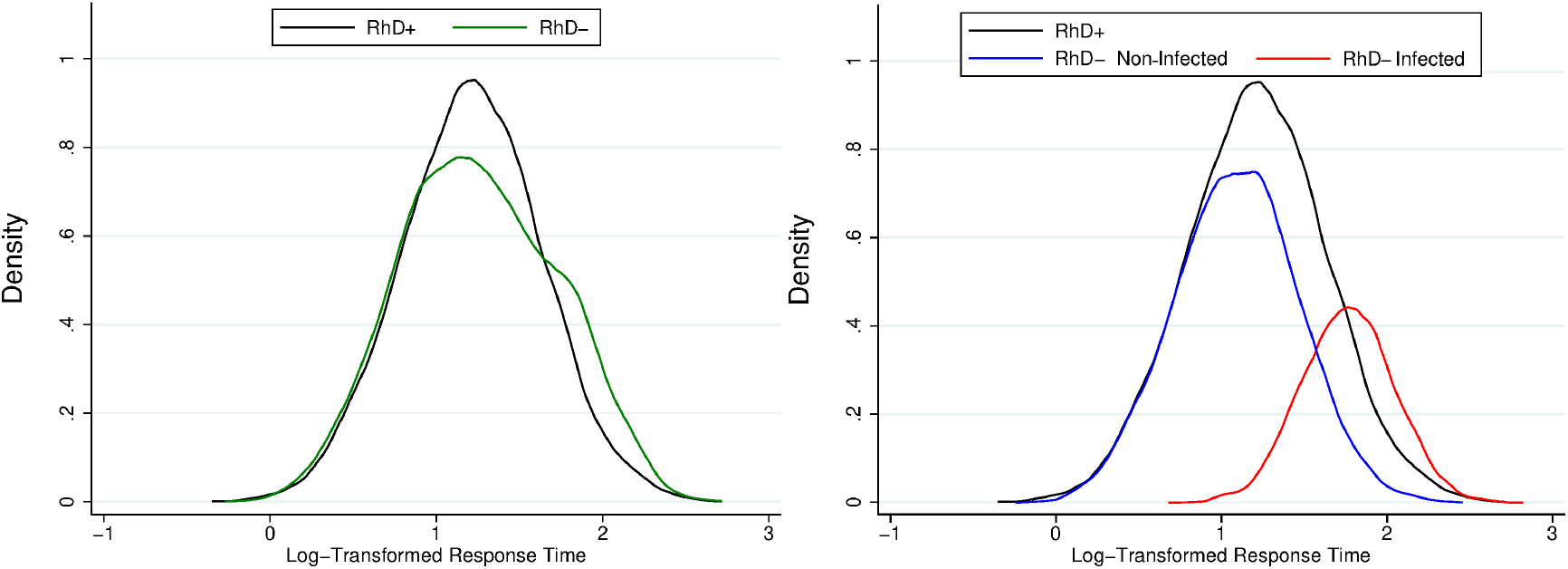
Left: Empirical distribution of log-transformed response times for the survey study (*N* = 2, 020) depending on blood type. Right: Applying a finite-mixture method, we recover the distributions for (imputed) infected and non-infected within the RhD-negative population, and estimate the extent of the infection at 18.32%.

All results below refer to RhD-negative participants, for which the classification based on response times allows us to establish an imputed infection status. We remark that, to further establish that the classification from our method is responsive to Toxoplasmosis-induced physiological changes, the survey included equal numbers of RhD-negative and positive participants. We applied the same method to RhD-positive participants, where Toxoplasmosis should not lead to noticeable physiological changes in response times. The method mechanically creates two groups, but, as expected, that classification does not produce any statistically significant difference between “afflicted” and “non-afflicted” RhD-positive participants in any of the tasks included in the survey.

All tests in the remainder of the article are between subjects. We report mean differences and 95% confidence intervals, but the reported *p*-values correspond to non-parametric tests (Mann-Whitney-Wilcoxon). Figures 4 and 5 further report effect sizes (as the difference in means divided by standard deviations). The Online Appendix contains the complete list of questions and tests for both studies.

**Figure 4:**
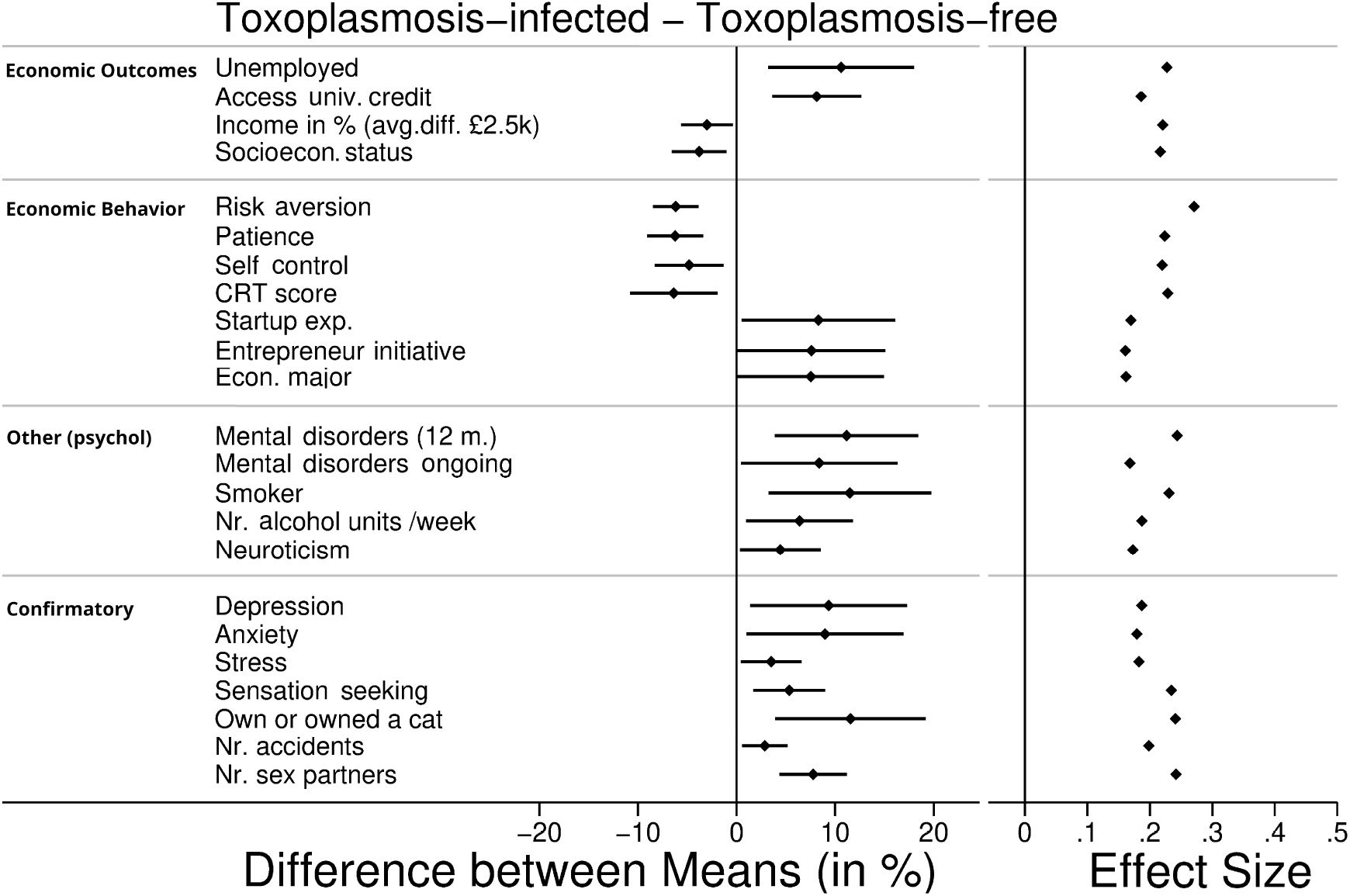
Differences between RhD-negative participants classified as infected and non-infected by the RT-method in a representative UK survey (*N* = 2, 020). We report differences between the means of the two groups (with 95% confidence intervals) and effect sizes. The upper-most part of the figure presents results on economic outcomes. The second block reports results on economic behavior. The third presents new results on mental health and addiction. As an additional validation of the method, the lower part confirms previously-reported effects of Toxoplasmosis while using our imputed classification.

**Figure 5:**
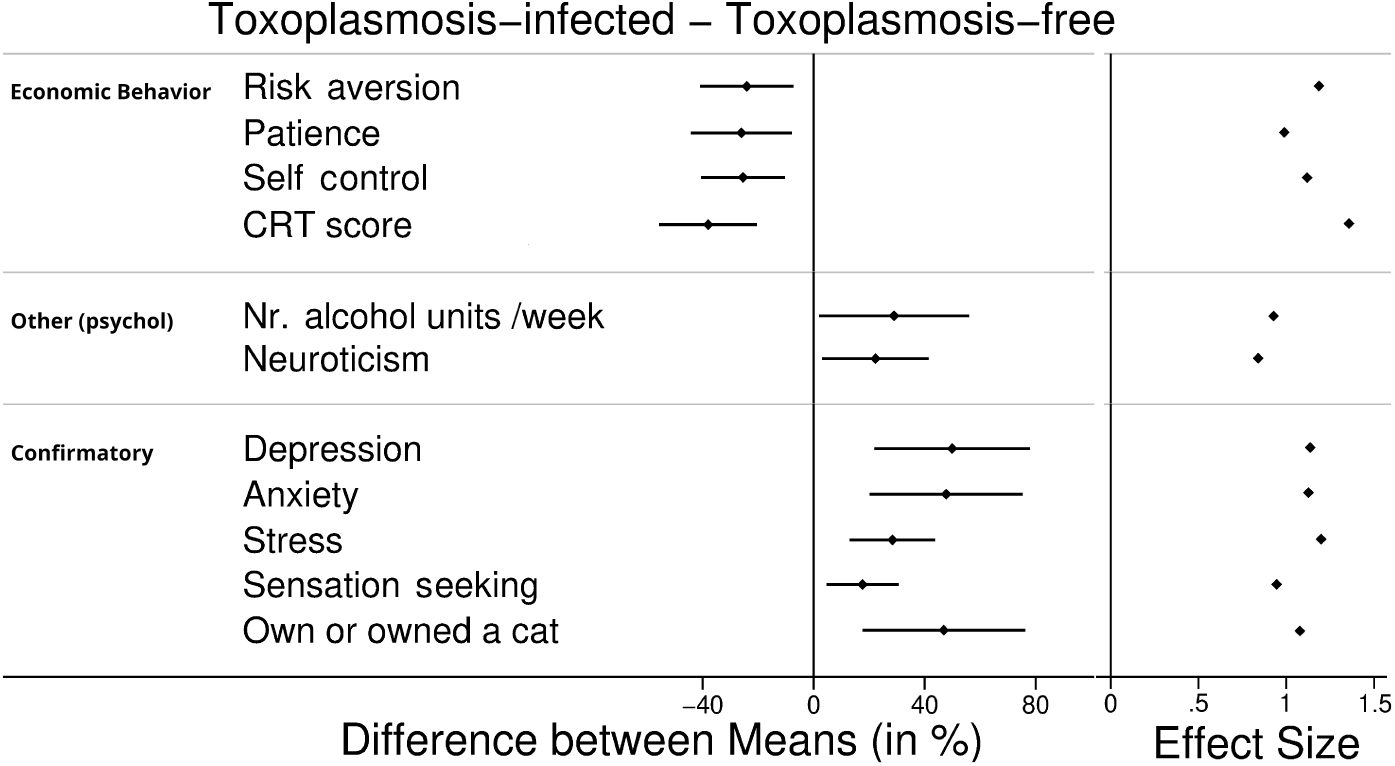
Differences between Toxoplasmosis-infected and Toxoplasmosis-free subjects in the clinical study (*N* = 79), where infection status is based on Toxoplasma IgG seroprevalence. We report differences between the means of the two groups (with 95% confidence intervals) and effect sizes (means difference divided by standard deviations). The upper part of the figure presents results on economic behavior. The middle part reports new results on mental health and addiction. The lower part confirms previously-reported effects of Toxoplasmosis.

### A Results: Economic Variables

Previous research on the behavioral and socioeconomic effects of Toxoplasmosis has been limited because, for large samples, current medical tests are expensive and time-consuming. Using self-reports is less than desirable because Toxoplasmosis infections rarely produce acute symptoms, and making subjects aware of the research topic might introduce sampling biases or experimenter demand effects. The proposed method is hence a substantial improvement over existing procedures to investigate economic and behavioral effects of Toxoplasmosis. Applying it to our data, we find several effects which are new to the literature.

Possible economic effects of the infection might be related to the disruption of dopamine regulation by the parasite. Dopamine disruption makes mice (the parasite’s target) less afraid and more willing to explore, and hence increase their chance of being eaten by cats (Vyas et al., 2007), closing the parasite’s life cycle. Due to the similarities between human and mice brains, Toxoplasmosis induces similar dopamine dis-regulations in humans. Infected subjects might hence become more impulsive, sensation-seeking, impatient, and risk-seeking. This could result in selecting riskier jobs (leading to higher unemployment), having more accidents, and so on, ultimately leading to lower yearly income and socioeconomic status.

Our data confirms these hypotheses (upper-most block in Figure 4). For this purpose, we make use of the fact that Prolific-registered participants provide answers to a number of standardized questions when registering for participation. We find clear, negative effects of the infection on economic outcomes. Toxoplasmosis-infected show a higher probability of being unemployed at the moment (mean difference 0.106; CI= [0.032, 0.180]; *p* = 0.0052). That is, in our survey, Toxoplasmosis is associated with a 10.6% increase in the probability of being unemployed. In agreement with this, Toxoplasmosis-infected are also more likely to have access to Universal Credit, a U.K. social security payment for those unemployed, unable to work, or earning below a poverty line (mean difference 0.081; CI= [0.011, 0.151]; *p* = 0.0228). Imputed-infected also report significantly lower income (mean difference −2547.641 GBP; CI= [326.160, 4769.123]; *p* = 0.0138). That is, in our survey, Toxoplasmosis is associated with an average reduction of around 2500 GBP in yearly income. Consistent with this observation, Toxoplasmosis-infected also report lower perceived socioeconomic status (mean difference −0.378; CI= [−0.655, −0.101]; *p* = 0.0085).

### B Results: Economic Behavior

Since the survey study is much larger than the clinical trial (*N* = 1, 010 vs. *N* = 79 RhD-negatives) and comprises a general-population sample instead of a homogeneous population of university students, it allows us to also study behavioral which are difficult to explore in small samples. We included a number of questions of interest for economic behavior (e.g., risk, patience, self-control) both in the survey (second block in Figure 4) and in the clinical trial (upper-most block in Figure 5).

In particular, we included the survey questions on risk attitudes and intertemporal discounting (patience) from Falk et al. (2018). We find that Toxoplasmosis-infected exhibit lower risk aversion, both in the survey study (mean difference −0.616; CI= [−0.978, −0.255]; *p* = 0.0007) and in the clinical study (−3.052; CI= [−4.759, −1.346]; *p* = 0.0004). They also exhibit lower patience, again both in the survey study (mean difference −0.627; CI= [−1.067, −0.180]; *p* = 0.0084) and in the clinical study (−2.342; CI= [−4.003, −0.681]; *p* = 0.0033).

To examine differences in self-control, we used the standard self-control scale of Tangney et al. (2004). We find that Toxoplasmosis-infected have lower reported self-control scores, both in the survey study (mean difference −2.602; CI= [−4.486, −0.718]; *p* = 0.0077) and in the clinical study (−12.714; CI= [−20.391, −5.036]; *p* = 0.0010). Our questions also included the Cognitive Reflection Test of Frederick (2005), in the extended version of Toplak et al. (2014) (where scores range from 0 to 7), which can be seen as a proxy for reflective vs. impulsive thinking. Toxoplasmosis-infected had worse Cognitive Reflection Test scores, both in the survey study (mean difference −0.446; CI= [0.136, 0.756]; *p* = 0.0044) and in the clinical study (−2.690; CI= [−3.942, −1.437]; *p* = 0.0001).

Reflecting the possible increase in risky and explorative behaviors among the infected, the imputed-infected in our survey study are more likely to have experience with startups (mean difference 0.083; CI= [0.005, 0.161]; *p* = 0.0370) and to have engaged in entrepreneurship (Johnson et al., 2018) (mean difference 0.075; CI= [0.000, 0.151]; *p* = 0.0481). Interestingly, they are also more likely to hold an economics- or management-related major (mean difference 0.075; CI= [0.001, 0.150]; *p* = 0.0466).^3^

### C Other New Results

Additional standard questions from Prolific allow us to establish further associations in the survey study (third block in Figure 4). In particular, related to the neuropsychiatric conditions associated with Toxoplasmosis (Sutterland et al., 2015), we also observe that Toxoplasmosis-infected are more likely to report having experienced a mental health disorder in the last 12 months (0.111; CI= [0.039.0.184]; *p* = 0.0027) and also more likely to have an ongoing mental health disorder (0.083; CI= [0.004, 0.163]; *p* = 0.0386).

In line with the lower self-control and higher sensation seeking of Toxoplasmosis-infected participants, we also observe that they are more likely to be smokers (mean difference 0.115; CI= [0.032, 0.198]; *p* = 0.0066). Our own survey questions (used in both studies; third block in Figure 4 and second block in Figure 5) included alcohol consumption, and we observe that Toxoplasmosis-infected consume a larger number of alcohol units per week than Toxoplasmosis-free subjects (survey study, mean difference 1.272; CI= [0.189, 2.357]; *p* = 0.0114; clinical study, 7.139; CI= [1.643, 12.636]; *p* = 0.0104).

Last, Toxoplasmosis-infected exhibited higher neuroticism scores as measured by the reduced Big-5 questionnaire of Gosling et al. (2003), which we included both in the survey study (mean difference 0.576; CI= [0.045.1.107]; *p* = 0.0426) and the clinical study (2.674; CI = [0.334, 5.014]; *p* = 0.0248).

## IV Confirmatory Results and Additional Validation

In both of our studies (clinical trial and survey studies), we included a number of questions for validation purposes. Those are related to documented effects of Toxoplasmosis, e.g. the relation to stress, anxiety, or sensation-seeking behavior.

### A Clinical Study

In the clinical trial (last block in Figure 5), we find that Toxoplasmosis-infected subjects are more likely to experience depression (Nayeri Chegeni et al., 2019) (mean difference 0.499; 95% CI= [0.215, 0.783]; *p* = 0.0037), anxiety (Flegr and Kaňková, 2022) (0.485; CI= [0.205, 0.764]; *p* = 0.0020), and stress (Flegr and Kaňková, 2022) (0.283; CI= [0.127, 0.440]; *p* = 0.0004), and are more sensation seeking (Cook et al., 2015) (1.757; CI= [0.436, 3.079]; *p* = 0.0081), compared to Toxoplasmosis-free ones. Since the comparison is based on infection status according to a medical test but for a small sample, these results are merely a replication of known effects. However, the results are qualitatively identical if we use the response-time classification.

### B Survey Study

In the survey study, with a far larger sample size, questions related to known effects of Toxoplasmosis served the additional purpose of showing that individuals classified as imputed-infected by our RT-method display the same psychological and neuropsychiatric alterations as those established to be infected by medical tests, hence providing an indirect, supplementary validation of the classification method.

Results are shown in the last block in Figure 4. In agreement with the literature, we find that imputed-infected survey participants are more likely to experience depression (Nayeri Chegeni et al., 2019) (mean difference 0.094; CI= [0.014, 0.173]; *p* = 0.0212), anxiety (Flegr and Kaňková, 2022) (0.090; CI= [0.010, 0.170]; *p* = 0.0272), and stress (Flegr and Kaňková, 2022) (0.035; CI= [0.005, 0.066]; *p* = 0.0240), and are more sensation seeking (Cook et al., 2015) (0.536; CI= [0.173, 0.900]; *p* = 0.0062), compared to imputed-non-infected participants. This reproduces known effects from the literature for our imputed infection status, hence indirectly validating the method. The results fully agree with those of our clinical trial, where the classification was based on a blood test (Figure 5).

We also observe that participants classified as infected are more likely to own or having owned a cat (0.116; CI= [0.040, 0.192]; *p* = 0.0030), an effect we also observed in our clinical study (0.469; CI= [0.171, 0.766]; *p* = 0.0050). Felines are the main vector of contagion for Toxoplasmosis as they are the only known definitive host for the parasite. They shed up to millions of oocysts per day in their feces, which sporulate and become infective in the environment (Kaushik et al., 2014). Hence, this is a further indirect validation of our classification.

In agreement with the literature (Flegr et al., 2002), in our survey study we also find that imputed-infected experienced a higher number of car accidents (0.344; CI= [0.069, 0.620]; *p* = 0.0236). Also, imputed-infected report a higher number of sexual partners (1.948; CI= [0.620, 3.275]; *p* = 0.0114), in agreement with previous results suggesting that the infected are viewed as more attractive (Borráz-León et al., 2022). However, these two questions did not lead to significant differences in the small student sample in our clinical study.

## V Discussion

We propose an inexpensive method to measure the prevalence of latent Toxoplasmosis infection, and validate it in a clinical trial. We then use this method to study the economic and behavioral effects of the infection in a large general-population sample.

The first benefit of the new method is its extremely low cost and ease of application, as it relies only on the response times from a three-minutes task. The test is of course less precise than an antibodies test, but still remarkably accurate. It can hence be used for large-scale testing even in low-income regions. Large-scale deployment can then be an efficient, complementary instrument to inform policy makers, identify most-affected subgroups, and adequately allocate scarce resources. This will help to reduce, prevent, and eventually eradicate the infection.

The main benefit of the method is to allow for systematic, large-scale studies of the (economic) consequences of latent Toxoplasmosis infections. The method is an easy-to-implement proxy for the medical test, and it can be deployed even in online surveys. We applied it to a general-population sample in the UK and found widespread and pervasive economic, behavioral, and psychological effects of latent infections. This is important because the literature has typically focused on acute infections, but our method establishes that the consequences of latent Toxoplasmosis infections are relevant and pervasive.

A limitation of our method is that we leverage physiological differences in the distribution of response times between infected and non-infected which depend on the Rhesus blood factor. Hence, the method requires this information. However, knowledge of blood type is common and tests for it are much faster and cheaper than the antigen tests required for the diagnosis of Toxoplasmosis.

The reported results on economic behavior and outcomes are in line with a common mechanism leading to behavioural changes in Toxoplasmosis-infected: the disruption of dopamine by the parasite. Due to the central role of dopamine in the human brain’s reward valuation network, its disruption has been shown to relate to mental disorders, depression, stress, and anxiety. Dopamine also regulates fear and anxiety (de la Mora et al., 2010; Abraham et al., 2014) and hence the disruption makes mice (the parasite’s target) less fearful and more willing to explore. In turn, this increase the chances that the mice will be eaten by cats (Vyas et al., 2007; Afonso et al., 2012), which closes the parasite’s life cycle. Toxoplasmosis seems to induce similar dopamine disregulations in humans, making infected subjects become more impulsive, sensation-seeking, and, in terms of economic behavior, also more impatient, and risk-seeking. These behavioral alternations can result in selecting riskier jobs and entering riskier economic activities (including startups and entrepreneurial initiatives), leading to higher unemployment, increased accidents, and other negative outcomes. Overall, the parasite-induced alterations lead to lower yearly income and decreased socioeconomic status. Given the large number of latent infections in the world population, these economic effects can be quite substantial in the aggregate.

## Data Availability

Raw data files for each study and the code for the analyses are available on OSF.

https://osf.io/j2rku/?view_only=482ca01c948044a08f5a8244172d4090

## Footnotes

1 The procedures used a Chemiluminescence Microparticle Immunoassay on the Architect i1000SR (Abbott) Analyzer and a u-capture ELISA for confirmation of reactive Architect samples. Serum IgG antibodies for Toxoplasma gondii usually peak in 2 to 3 months and then gradually decline to a lower but still detectable level characteristic of a chronic infection (Robert-Gangneux and Dardé, 2012). The cutoff threshold for positivity was set to 3.0 (IU/ml) following standards. The test is accredited according to the ISO 17025 standard and has a diagnostic specificity of 98.45% and sensitivity of 98.25%.

2 Prolific recruits representative samples and applies the required filters afterwards, e.g. having reported the blood type. In our case, this resulted in an oversampling of females. Other variables were unaffected, e.g. the median age in our study was 40 years, compared to the UK’s latest-released median age of 40.7 (Office for National Statistics (ONS),

3 Among all participants who reported having started studies related to economics, management, business, finance, or commerce, the percentage of imputed-infected was 21.88%, compared to 16.67% for other participants. The difference is significant according to a test of proportions (two-sided; *p* = 0.0465). 2024).

