## Supplementary material for "Two Billion Infected: An Inexpensive Method to Measure Latent Toxoplasmosis and its Economic Consequences^*^": Online Appendix

#### 1 Complete List of Tasks and Tests

In this section we report the complete list of statistical tests which, for reasons of brevity, could not be listed in the main text of the paper. All tests report the difference between mean behavior of infected vs. non-infected, confidence intervals for the corresponding $t$-test, and $p$-values for the respective non-parametric Mann-Whitney U Test.

Clinical-trial study:

- Owning or having owned a cat: 0.469; 95% CI= $[0.171, 0.766]$, $p = 0.0050$.
- Unemployment: 0.153; 95% CI= $[-0.458, 0.152]$, $p = 0.5031$.
- Experience with entrepreneurship: 0.121; 95% CI= $[-0.030, 0.273]$, $p = 0.3268$.
- Experience with startup: 0.117; 95% CI= $[-0.161, 0.395]$, $p = 0.6495$.
- Self-reported being neurodivergent: 0.041; 95% CI= $[-0.219, 0.300]$, $p = 1.0000$.
- Number of alcohol units consumed weekly: 7.139; 95% CI= $[1.643, 12.636]$, $p = 0.0104$.
- Self-reported suffering from mental disorders in the last 12 months: 0.124; 95% CI= $[-0.139, 0.388]$, $p = 0.5465$.
- Self-reported ongoing mental disorders: -0.051; 95% CI= $[-0.261, 0.159]$, $p = 1.0000$.
- Suffering from depression: 0.499; 95% CI= $[0.215, 0.783]$, $p = 0.0037$.
- Suffering from anxiety: 0.485; 95% CI= $[0.205, 0.764]$, $p = 0.0020$.
- Suffering from ADHD: 0.116; 95% CI= $[0.110, 0.341]$, $p = 0.5243$.
- Smoking (current or past frequent): -0.036; 95% CI= $[-0.237, 0.165]$, $p = 1.0000$.

*Brain, Mind & Markets Laboratory, University of Melbourne, Melbourne, Australia.
†Department of Political and Social Sciences, Zeppelin University Friedrichshafen, Germany.

- Income: -5475.124; 95% CI= $[-15695.700, 4745.455]$, $p = 0.1370$.
- University major in an economic topic: 0.047; 95% CI= $[-0.163, 0.257]$, $p = 0.9434$.
- Number of sexual partners: -0.034; 95% CI= $[-3.018, 2.950]$, $p = 0.4491$.
- Left-handedness (used as a robustness test that the classification based on RTs does not produce artificial differences): -0.090; 95% CI= $[-0.256, 0.077]$, $p = 0.7181$.
- Saving for retirement: -0.081; 95% CI= $[-0.307, 0.145]$, $p = 0.8365$.
- Owning investments: -0.123; 95% CI= $[-0.424, 0.178]$, $p = 0.6362$.
- Risk attitudes: -3.052; 95% CI= $[-4.759, -1.346]$, $p = 0.0004$.
- Number of car accidents: 0.019; 95% CI= $[-0.668, 0.706]$, $p = 0.9694$.
- Number of accidents at work: 0.731; 95% CI= $[-0.102, 1.564]$, $p = 0.7381$. This is significant according to a $t$-test (p=0.0422).
- CRT score: -2.690; 95% CI= $[-3.942, -1.437]$, $p = 0.0001$.
- Stress: 0.283; 95% CI= $[0.127, 0.440]$, $p = 0.0004$.
- Score in the self-control scale: -12.714; 95% CI= $[-20.391, -5.036]$, $p = 0.0010$.
- Extraversion: 0.430; 95% CI= $[-1.691, 2.552]$, $p = 0.7528$.
- Agreeableness: 0.606; 95% CI= $[-0.709, 1.920]$, $p = 0.5082$.
- Conscientiousness: 0.568; 95% CI= $[-0.875, 2.012]$, $p = 0.2933$.
- Neuroticism (stability): 2.674; 95% CI= $[0.334, 5.014]$, $p = 0.0248$.
- Openness: -1.004; 95% CI= $[-2.221, 0.214]$, $p = 0.1284$. This is significant according to a $t$-test (p=0.0524).
- Sensation seeking score: 1.757; 95% CI= $[0.436, 3.079]$, $p = 0.0081$.
- Patience: -2.342; 95% CI= $[-4.003, -0.681]$, $p = 0.0033$.

Survey study:

- Owning or having owned a cat: 0.116; 95% CI= $[0.040, 0.192]$, $p = 0.0030$.
- Unemployment: 0.106; 95% CI= $[0.032, 0.180]$, $p = 0.0052$.
- Experience with entrepreneurship: 0.075; 95% CI= $[0.000, 0.151]$, $p = 0.0481$.
- Experience with startup: 0.083; 95% CI= $[0.005, 0.161]$, $p = 0.0370$.
- University degree: 0.018; 95% CI= $[-0.053, 0.090]$, $p = 0.6142$.
- Self-reported being neurodivergent: -0.016; 95% CI= $[-0.081, 0.049]$, $p = 0.6294$.
- Number of alcohol units consumed weekly: 1.272; 95% CI= $[0.189, 2.357]$, $p = 0.0114$.
- Self-reported suffering from mental disorders in the last 12 months: 0.111; 95% CI= $[0.039, 0.184]$, $p = 0.0027$.

- Self-reported ongoing mental disorders: 0.083; 95% CI= $[0.004, 0.163]$, $p = 0.0386$.
- Suffering from depression: 0.094; 95% CI= $[0.014, 0.173]$, $p = 0.0212$.
- Suffering from anxiety: 0.090; 95% CI= $[0.010, 0.170]$, $p = 0.0272$.
- Suffering from ADHD: -0.008; 95% CI= $[-0.057, 0.041]$, $p = 0.7515$.
- Smoking (current or past frequent): 0.115; 95% CI= $[0.032, 0.198]$, $p = 0.0066$.
- Income: -2547.641; 95% CI= $[326.160, 4769.123]$, $p = 0.0138$.
- University major in an economic topic: 0.075; 95% CI= $[0.001, 0.150]$, $p = 0.0466$.
- Self-Reported Socioeconomic status (ladder): -0.378; 95% CI= $[-0.655, -0.101]$, $p = 0.0085$.
- Number of sexual partners: 1.948; 95% CI= $[0.620, 3.275]$, $p = 0.011$.
- Left-handedness (used as a robustness that the classification based on RTs does not produce artificial differences): 0.000; 95% CI= $[-0.050, 0.051]$, $p = 0.9869$.
- Saving for retirement: 0.076; 95% CI= $[-0.003, 0.155]$, $p = 0.0616$. This is significant according to a $t$-test ($p = 0.0308$).
- Owning investments: 0.082; 95% CI= $[0.002, 0.163]$, $p = 0.0444$.
- Risk attitudes: -0.616; 95% CI= $[-0.978, -0.255]$, $p = 0.0007$.
- Number of car accidents: 0.344; 95% CI= $[0.069, 0.620]$, $p = 0.0236$.
- Number of accidents at work: 0.054; 95% CI= $[-0.134, 0.242]$, $p = 0.0381$. This is not statistically significant according to a $t$-test, which is why the CIs cross the 0.
- CRT score: -0.446; 95% CI= $[-0.756, -0.136]$, $p = 0.0044$.
- Stress: 0.035; 95% CI= $[0.005, 0.066]$, $p = 0.0240$.
- Score in the self-control scale: -2.602; 95% CI= $[-4.486, -0.718]$, $p = 0.0077$.
- Extraversion: -0.223; 95% CI= $[-0.731, 0.285]$, $p = 0.4825$.
- Agreeableness: 0.206; 95% CI= $[-0.163, 0.576]$, $p = 0.3407$.
- Conscientiousness: 0.012; 95% CI= $[-0.397, 0.420]$, $p = 0.9508$.
- Neuroticism (stability): 0.576; 95% CI= $[0.045, 1.107]$, $p = 0.0426$.
- Openness: -0.303; 95% CI= $[-0.681, 0.076]$, $p = 0.0546$.
- Sensation seeking score: 0.536; 95% CI= $[0.173, 0.900]$, $p = 0.0062$.
- Patience: -0.627; 95% CI= $[-1.067, -0.180]$, $p = 0.008$.

### 2 Instructions

#### 2.1 Prolific Questions

The following questions are part of the Prolific questionnaire participants answered before enrolling into our survey study. To ensure comparability, we kept the same wording for these questions in the clinical-trial study whenever possible. Replies to these questions were voluntary. Participants were encouraged by Prolific to answer these questions as doing so increases the probability of receiving invitations to participate into further studies which require some specific sub-populations.

- What is your employment status?
  - Full-Time
  - Part-Time
  - Due to start a new job within the next month
  - Unemployed (and job seeking)
  - Not in paid work (e.g. homemaker, retired or disabled)

  Full-time, part-time, and "Not in paid work" are classified as employed.

- Have you engaged in entrepreneurship/run your own business?
  - I have in the past
  - I am currently doing this
  - I intend to in the future

  The first two options are classified as a yes.

- Have you ever considered a career move as a (paid) employee in a startup?
  - Yes, I'm doing it now
  - Yes, I've done it in the past
  - Yes, I've applied but it didn't happen
  - Yes, I've considered it
  - Yes, I've looked into seriously
  - No, I've never considered it.

  The first three options are classified as a yes.

- Which of these is the highest level of education you have completed? [Not asked in the clinical study]
  - No formal qualifications
  - Secondary education (e.g. GED/GCSE)
  - High school diploma/A-levels
  - Technical/community college
  - Undergraduate degree (BA/BSc/other)
  - Graduate degree (MA/MSc/MPhil/other)

  - Doctorate degree (PhD/other)

  The first three options are classified as a no.

- Do you consider yourself to be neurodivergent?
  - Yes
  - No
- What is your blood type?
  - A RhD positive (A+)
  - A RhD negative (A-)
  - B RhD positive (B+)
  - B RhD negative (B-)
  - O RhD positive (O+)
  - O RhD negative (O-)
  - AB RhD positive (AB+)
  - AB RhD negative (AB-)
- How many units of alcohol do you drink on average per week?
  - 0
  - 1-4
  - 5-9
  - 10-13
  - 14+
- Have you tried to access mental health support on the NHS in the last 12 months? [omitted "on the NHS" in the clinical study]
  - Yes
  - No
- Do you have – or have you had – a diagnosed, on-going mental health/illness/condition?
  - Yes
  - No
- Do you experience depression?
  - Yes
  - No
- Do you experience anxiety?
  - Yes
  - No
- Do you consider yourself to have attention deficit disorder (ADD)/attention deficit hyperactivity disorder (ADHD)?

  - Yes
  - No

- What is your current smoking status? Please select from the following options:
  - I am a current smoker (smoke at least 5 cigarettes a day and have smoked this amount for at least one year)
  - I am a recent smoker (smoke at least 5 cigarettes a day and have smoked this amount for less than one year)
  - I am a former smoker (used to smoke at least 5 cigarettes a day and smoked this amount for at least one year)
  - I have never smoked (smoked fewer than 100 cigarettes in my lifetime)

  The first three options are classified as a yes.

- How many romantic partners have you had? These can be short-term (casual, one night stands, friends with benefits etc.), or long-term (committed relationships). [Open field in the clinical study]
  - 0
  - 1
  - 2
  - 3
  - 4
  - 5
  - 6
  - 7
  - 8
  - 9
  - 10
  - 11-15
  - 16-20
  - More than 21

- Do you currently own any of the following as a pet?
  - Dog
  - Cat
  - Fish
  - Bird
  - Rabbit
  - Reptile
  - Other small mammal (e.g. hamster)
  - Do not have a pet

- Are you left or right-handed?

  - Right-handed
  - Left-handed
  - Ambidextrous

  Ambidextrous are classified as left-handed.

- Where would you put yourself on the socioeconomic ladder? Think of a ladder as representing where people stand in society. At the top of the ladder are the people who are best off—those who have the most money, most education and the best jobs. At the bottom are the people who are worst off—who have the least money, least education and the worst jobs or no job. The higher up you are on this ladder, the closer you are to people at the very top and the lower you are, the closer you are to the bottom. Choose the number whose position best represents where you would be on this ladder.

  [Scale of one to ten]

- Do you have a retirement plan?
  - Yes
  - No

- Have you ever made investments (either personal or through your employment) in the common stock or shares of a company?
  - Yes
  - No

- What is your personal income per year (after tax) in GBP? [Open field in the clinical study]
  - Less than £10,000
  - £10,000 - £19,999
  - £20,000 - £29,999
  - £30,000 - £39,999
  - £40,000 - £49,999
  - £50,000 - £59,999
  - £60,000 - £69,999
  - £70,000 - £79,999
  - £80,000 - £89,999
  - £90,000 - £99,999
  - £100,000 - £149,999
  - More than £150,000

- Are you a recipient of Universal Credit? [Not asked in the clinical study]
  - Yes
  - No

##### 2.2 Main Questions

Parts in [ ] are not displayed to participants.

- Reaction-time task from Havlíček et al. (2001); Novotná et al. (2008):

  In this part of the experiment you will be asked to perform a reaction time task. In the next screens the "Continue" button will appear in the centre of the display at irregular intervals. Your task is to click on the "Continue" button as quickly as possible.

  This task will last 3 minutes. Please try to keep your attention for the entire duration of the task.

  [A field appears in the center of the screen at irregular intervals ranging from 1 to 8 sec. The subjects have to respond to the square immediately after it appeared by pressing on it.]

- Version of the Big-5 personality trait from Gosling et al. (2003):

  Here are a number of personality traits that may or may not apply to you. Please indicate a number for each statement to indicate the extent to which you agree or disagree with that statement. You should rate the extent to which the pair of traits applies to you, even if one characteristic applies more strongly than the other.

  I see myself as:

  - Extraverted, enthusiastic: [7 point Likert scale with the following labels: Disagree strongly, Disagree moderately, Disagree a little, Neither agree not disagree, Agree a little, Agree moderately, Agree strongly]
  - Critical, quarrelsome: [7 point Likert scale with the following labels: Disagree strongly, Disagree moderately, Disagree a little, Neither agree not disagree, Agree a little, Agree moderately, Agree strongly]
  - Dependable, self-disciplined: [7 point Likert scale with the following labels: Disagree strongly, Disagree moderately, Disagree a little, Neither agree not disagree, Agree a little, Agree moderately, Agree strongly]
  - Anxious, easily upset: [7 point Likert scale with the following labels: Disagree strongly, Disagree moderately, Disagree a little, Neither agree not disagree, Agree a little, Agree moderately, Agree strongly]
  - Open to new experiences, complex: [7 point Likert scale with the following labels: Disagree strongly, Disagree moderately, Disagree a little, Neither agree not disagree, Agree a little, Agree moderately, Agree strongly]
  - Reserved, quiet: [7 point Likert scale with the following labels: Disagree strongly, Disagree moderately, Disagree a little, Neither agree not disagree, Agree a little, Agree moderately, Agree strongly]
  - Sympathetic, warm: [7 point Likert scale with the following labels: Disagree strongly, Disagree moderately, Disagree a little, Neither agree not disagree, Agree a little, Agree moderately, Agree strongly]
  - Disorganized, careless: [7 point Likert scale with the following labels: Disagree strongly, Disagree moderately, Disagree a little, Neither agree not disagree, Agree a little, Agree moderately, Agree strongly]

- Calm, emotionally stable: [7 point Likert scale with the following labels: Disagree strongly, Disagree moderately, Disagree a little, Neither agree not disagree, Agree a little, Agree moderately, Agree strongly]
- Conventional, uncreative: [7 point Likert scale with the following labels: Disagree strongly, Disagree moderately, Disagree a little, Neither agree not disagree, Agree a little, Agree moderately, Agree strongly]

- Experience Seeking Scale from Cook et al. (2015):

  Each of the questions below contains two choices. Please indicate the choice which most describes the way you feel. In some cases you may find items in which both choices describe your likes or feelings. Please choose the one which better describes your likes or feelings. In some cases you may find items in which you do not like either choice. In these cases mark the choice you dislike least. We are interested only in your feelings, not in how others feel about these things or how one is supposed to feel. There are not right or wrong answers as in other kinds of tests.

  - Question 1: I dislike body odors/I like some of the earthy body smells.
  - Question 2: I like to explore a strange city or section of town by myself, even if it means getting lost/I prefer a guide when I am in a place I don't know well.
  - Question 3: I have tried marijuana or would like to./ I would never smoke marijuana.
  - Question 4: I would not like to try any drug which produces strange and dangerous effects on me./ I would like to try some of the new drugs that produce hallucinations.
  - Question 5: I like to try new foods that I have never tasted before./I order the dishes with which I am familiar, so to avoid disappointment and unpleasantness.
  - Question 6: I would like to take off on a trip with no pre-planned or definite routes, or timetable./When I go on a trip I like to plan my route and timetable fairly carefully.
  - Question 7: I prefer the "down-to-earth" kinds of people as friends./ I would like to make friends in some of the "far-out" groups like artists or "punks".
  - Question 8: A sensible person avoids activities that are dangerous./I sometimes like to do things that are a little frightening.
  - Question 9: The essence of good art is its clarity, symmetry of form and harmony of colors./I often find beauty in the "clashing" colors and irregular forms of modern paintings.
  - Question 10: People should dress according to some standards of taste, neatness, and style./People should dress in individual ways even if the effects are sometimes strange.

- Cognitive Reflection Test from Toplak et al. (2014):

  - A postcard and a pen cost 110 cents in total. The postcard costs 100 cents more than the pen. How much does the pen cost? (in cents)

  - If it takes 5 machines 5 minutes to make 5 car tires, how long would it take 100 machines to make 100 car tires?
  - In a lake, there is a patch of lily pads. Every day, the patch doubles in size. If it takes 48 days for the patch to cover the entire lake, how long would it take for the patch to cover half of the lake? (in days)
  - If John can drink one barrel of water in 6 days, and Mary can drink one barrel of water in 12 days, how long would it take them to drink one barrel of water together? (in days)
  - Jerry received both the 15th highest and the 15th lowest mark in the class. How many students are in the class?
  - A man buys a pig for $60, sells it for $70, buys it back for $80, and sells it finally for $90. How much has he made? (in dollars)
  - Simon decided to invest $8,000 in the stock market one day early in 2008. Six months after he invested, on July 17, the stocks he had purchased were down 50%. Fortunately for Simon, from July 17 to October 17, the stocks he had purchased went up 75%. At this point, Simon:
    * has broken even
    * is ahead of where he began
    * has lost money

- Risk Preferences and Patience from Falk et al. (2018):

  - How do you see yourself: Are you generally a person who is fully prepared to take risks or do you try to avoid risks?

    Please indicate an option on the scale, where the value 1 means: not willing to take risks and value 10 means: very willing to take risks. [10-point Likert scale]
  - In comparison to others, are you a person who is generally willing to give up something today in order to benefit from that in the future? Please assume there is no inflation, i.e., future prices are the same as today's prices.

    Please indicate an option on the scale, where the value 1 means: not willing to give up something today and value 10 means: very willing to give up something today. [10-point Likert scale]

- Self-Control Scale from Tangney et al. (2004):

  Using the scale provided, please indicate how much each of the following statements reflects how you typically are: [5-point Likert scale from "Not at all" to "Very Much"]

  - I am good at resisting temptation.
  - I have a hard time breaking bad habits.
  - I am lazy.
  - I say inappropriate things.
  - I never allow myself to lose control.
  - I do certain things that are bad for me, if they are fun.
  - People can count on me to keep on schedule.

- Getting up in the morning is hard for me.
- I have trouble saying no.
- I change my mind fairly often.
- I blurt out whatever is on my mind.
- People would describe me as impulsive.
- I refuse things that are bad for me.
- I spend too much money.
- I keep everything neat.
- I am self-indulgent at times.
- I wish I had more self-discipline.
- I am reliable.
- I get carried away by my feelings.
- I do many things on the spur of the moment.
- I don't keep secrets very well.
- People would say that I have iron self-discipline.
- I have worked or studied all night at the last minute.
- I'm not easily discouraged.
- I'd be better off if I stopped to think before acting.
- I engage in healthy practices.
- I eat healthy foods.
- Pleasure and fun sometimes keep me from getting work done.
- I have trouble concentrating.
- I am able to work effectively toward long-term goals.
- Sometimes I can't stop myself from doing something, even if I know it is wrong.
- I often act without thinking through all the alternatives.
- I lose my temper too easily.
- I often interrupt people.
- I sometimes drink or use drugs to excess.
- I am always on time.

• What do you study at the university? [Open field. Any answer which contained the characters "econ", "manag", "busin", "account", or "financ" is classified as an economic major.

• In how many traffic accidents have you been involved in your life? [Open field]

• In how many work-related accidents have you been involved in your life? [Open field]

• Stress scale from Cohen et al. (1994):

  The questions below ask you about your feelings and thoughts during the last month. In each case, you will be asked to indicate how often you felt or thought a certain way. Possible answers to all questions were: Never, Almost never, Sometimes, Fairly often, Very often.

- In the last month, how often have you been upset because of something that happened unexpectedly?
- In the last month, how often have you felt that you were unable to control the important things in your life?
- In the last month, how often have you felt nervous and “stressed”?
- In the last month, how often have you felt confident about your ability to handle your personal problems?
- In the last month, how often have you felt that things were going your way?
- In the last month, how often have you found that you could not cope with all the things that you had to do?
- In the last month, how often have you been able to control irritations in your life?
- In the last month, how often have you felt that you were on top of things?
- In the last month, how often have you been angered because of things that were outside of your control?
- In the last month, how often have you felt difficulties were piling up so high that you could not overcome them?
